# Diagnosis in Bytes: Comparing the Diagnostic Accuracy of Google and ChatGPT 3.5 as Diagnostic Support Tools

**DOI:** 10.1101/2023.11.10.23294668

**Authors:** Guilherme R Guimaraes, Caroline Santos Silva, Jean Carlos Z Contreras, Ricardo G Figueiredo, Uros - Grupo de Pesquisa, Ricardo B Tiraboschi, Cristiano M Gomes, Jose de Bessa

**Affiliations:** Universidade Estadual de Feira de Santana – UEFS; Collaborative Research Group (undergraduate students) – Medical School Universidade Estadual de Feira de Santana – UEFS; Universidade de São Paulo - USP

**Keywords:** Medical Informatics Applications, Artificial Intelligence, Diagnosis, Urology

## Abstract

**Objective:** Adopting digital technologies as diagnostic support tools in medicine is unquestionable. However, the accuracy in suggesting diagnoses remains controversial and underexplored. We aimed to evaluate and compare the diagnostic accuracy of two primary and accessible internet search tools: Google and ChatGPT 3.5.

**Method:** We used 60 clinical cases related to urological pathologies to evaluate both platforms. These cases were divided into two groups: one with common conditions (constructed from the most frequent symptoms, following EAU and UpToDate guidelines) and another with rare disorders - based on case reports published between 2022 and 2023 in Urology Case Reports. Each case was inputted into Google Search and ChatGPT 3.5, and the results were categorized as "correct diagnosis," "likely differential diagnosis," or "incorrect diagnosis." A team of researchers evaluated the responses blindly and randomly.

**Results:** In typical cases, Google achieved 53.3% accuracy, offering a likely differential diagnosis in 23.3% and errors in the rest. ChatGPT 3.5 exhibited superior performance, with 86.6% accuracy, and suggested a reasonable differential diagnosis in 13.3%, without mistakes. In rare cases, Google did not provide correct diagnoses but offered a likely differential diagnosis in 20%. ChatGPT 3.5 achieved 16.6% accuracy, with 50% differential diagnoses.

**Conclusion:** ChatGPT 3.5 demonstrated higher diagnostic accuracy than Google in both contexts. The platform showed acceptable accuracy in common cases; however, limitations in rare cases remained evident.

## INTRODUCTION

The advent of the first data processing machines in the 1940s piqued the interest of experts from various fields for their potential applications, and medicine was no exception (1). As early as 1959, Brodman and colleagues demonstrated that a trained computerized system could identify patterns in a group of symptoms reported by patients and suggest possible diagnoses, performing comparably to physicians receiving the same information (2). Since then, data analytics technologies and Artificial Intelligence (AI) have gained prominence in various medical fields, including public health, medical image analysis, clinical trials, and more (3).

The increasing capability of these tools to integrate information has allowed researchers to envision diagnostic applications far beyond what Broadman presented. It is now possible to input a patient’s symptoms into everyday search tools and receive a list of likely diagnoses (3,4). Such utility was already explored by Tang and Ng in 2006, who assessed the frequency of accurate diagnoses provided when specific symptoms of specific diseases were searched on Google, the leading internet search site (5).

In addition to search tools, new chatbots have been explored for this purpose, merging AI with messaging interfaces, such that recent initial studies have attempted to evaluate the diagnostic accuracy of these instruments (6). The most famous of these is the Generative Pre-Trained Transformer - ChatGPT 3.5, developed by OpenAI, whose operational model is not solely based on delivering pages with more terms matching the search, like traditional search tools, but instead on providing real-time AI-generated text based on the extensive database on which the AI was trained (7,8).

In this context of evolving information technologies and their increasingly pervasive integration in medicine, this study aimed to evaluate and compare the diagnostic accuracy for urological cases of two leading internet search tools: Google and ChatGPT 3.5.

## METHODS

This diagnostic evaluation study was conducted between April and June 2023, in which the diagnostic accuracy of the platforms Google and ChatGPT 3.5 was assessed. These tools were evaluated based on the responses to 60 clinical cases related to urological pathologies, divided into "common" and "rare" conditions. Questions were formulated in Portuguese Language.

In the first group, 30 descriptions summarized the typical clinical presentation of common urological diseases, written under the European Association of Urology and UpToDate guidelines. In the second group, the remaining 30 cases were based on reports published between 2022 and 2023 in Urology Case Reports, selected based on the typicality of their manifestations (Supplementary File 1). Questions requiring extensive evaluation for specific diagnoses were excluded.

Each clinical case was inputted into Google Search and ChatGPT 3.5, and the results were categorized as "correct," likely and plausible," and "incorrect" according to the blind and random judgment of a panel of 3 researchers. For Google Search, an incognito tab in the browser was used, with no linked account, to minimize any influence from previous search history, and the first three displayed results were considered for diagnostic categorization. For ChatGPT 3.5, a specific "account" was created to reduce the influence of prior searches.

The results were described in absolute numbers and percentages. The Chi-square test was used for proportion comparison, and the Kappa test was employed to assess agreement between the instruments. GraphPad Prism version 10.0.0 for Windows, GraphPad Software, Boston, Massachusetts, USA, was used for analysis and graphs.

## RESULTS

Both platforms showed promising results when dealing with more typical or "common" urological cases. Google Search demonstrated a diagnostic accuracy rate of 16 (53.3%). It correctly identified the disease in over half of the topics and provided likely differential diagnoses in seven more (23.3%). However, it failed to produce the correct response in the remaining cases.

In contrast, ChatGPT 3.5 outperformed Google, achieving a correct response in 26 (86.6%) cases and offering likely differential diagnoses in another four (13.3%) cases, with no incorrect diagnoses.

Regarding the rarer urological conditions, the performances of the two platforms significantly diverged. Google could not produce correct diagnoses but offered likely differential diagnoses in six (20%) cases. Google’s suggestions were incorrect in the remaining 24 (80%) cases. On the other hand, ChatGPT 3.5 showed moderate success in diagnosing rare conditions, with a correct diagnostic rate of five (16.6%). Impressively, it provided a likely differential diagnosis in half of the cases, 15 (50%). However, it should be noted that the platform made diagnostic errors in the remaining instances (Figure 1).

**Figure 1:**
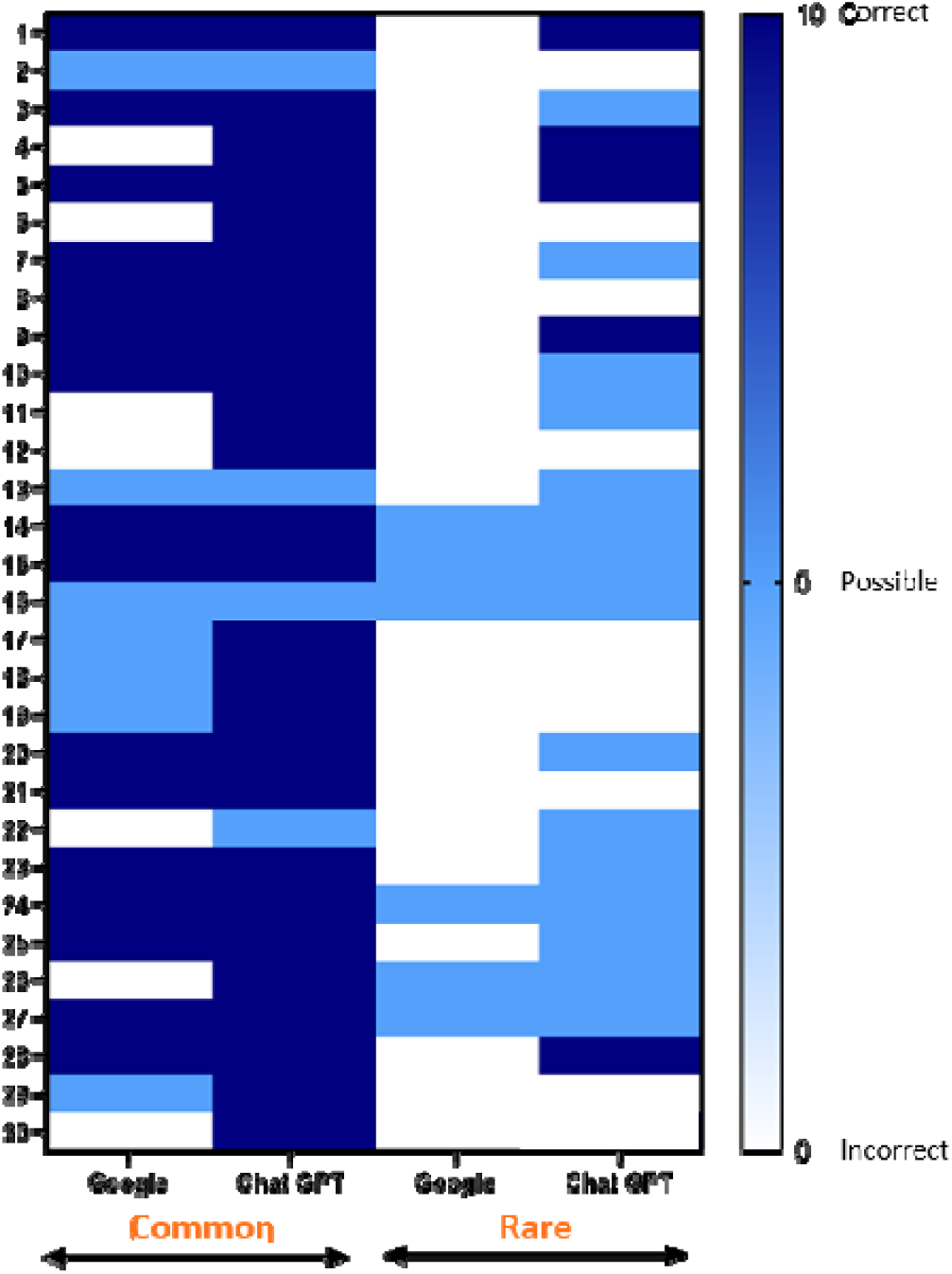
Comparison of Diagnostic Accuracy between Google and ChatGPT 3.5

When analyzed together, ChatGPT 3.5 was significantly superior to Google. It got it right or suggested an acceptable differential diagnosis in 50 (83.3%), while Google did so in 29 (48.3%). RR=1.46 p< 0.001. Both tools did not respond correctly to ten questions (rare cases). There was low agreement between the two tools. Kappa=0.315.

## DISCUSSION

We can demonstrate that ChatGPT 3.5 outperformed Google in this experiment, simulating real-life urological scenarios. Both platforms vary in performance depending on the complexity and rarity of urological conditions. While Google remained moderately effective in ordinary cases, its performance dropped significantly in the context of rarer diseases. On the other hand, ChatGPT 3.5 showed a high degree of accuracy in diagnosing common urological diseases and a moderate level of success in rare and uncommon conditions.

These findings indicate the need to acknowledge the variable performance of these tools in different clinical scenarios, highlighting their strengths and areas where improvements are needed. The results also raise essential questions about the role of AI-based platforms like ChatGPT 3.5 in clinical decision-making and education. The explosive growth of medical knowledge exacerbates the challenges faced by healthcare professionals. Estimates indicate that the time required for medical knowledge to double has drastically decreased - from 50 years in 1950 to 73 days in 2020(9). These rapid advances mean medical students must master 342 potential diagnoses for 37 frequently presented signs and symptoms before graduating(10). This volume of information can be overwhelming and underscores the growing need for accurate and efficient diagnostic tools to assist physicians and other healthcare providers.

General search engines like Google have existed for some time and have gradually found applications in the medical field(11,12). In 1999, Graber and colleagues assessed the use of search engines to answer medical questions, finding that these platforms could correctly answer half of the questions (11). By 2006, Google had already become the dominant search tool, providing correct diagnoses in 58% of cases in Tang and Ng’s study(5). Another study evaluated the diagnostic accuracy of medical students before and after consulting Google and PubMed, noting a statistically insignificant but interesting 9.9% increase in diagnostic accuracy(13).

More recently, chatbots, which combine AI with messaging interfaces, have emerged as precise tools for generating direct responses(14,15). A Japanese study showed that ChatGPT 3 achieved a correct diagnostic rate of 93.3% when considering a list of ten probable differential diagnoses(16). The same platform demonstrated an accuracy rate of 80% in questions about cirrhosis and hepatocellular carcinoma and an accuracy of 88% in breast cancer prevention and screening(17,18).

In the field of urology, ChatGPT 3.5’s performance has been variable. In one study, the platform correctly answered 92% of pediatric urology questions(19). However, the platform’s performance dropped to 52% in another assessment focused on general urology(20). As illustrated by Huynh and colleagues, ChatGPT 3.5 performed poorly in the American Urological Association’s 2022 self-assessment study program, scoring below 30%(21). Studies evaluating the quality of information provided to the general population about urological pathologies presented good information but not without bias(22,23).

The current study expands on existing research by directly comparing the diagnostic effectiveness of Google and ChatGPT 3.5 for urological conditions. Our results indicate that ChatGPT 3.5 outperformed Google in common and rare cases. While the platform showed high precision in diagnosing common urological conditions, it demonstrated moderate success with rarer diseases. These findings offer promising perspectives for integrating such tools in educational and clinical workflows.

These platforms can assist urological researchers in analyzing and interpreting data, writing articles, and creating educational material. Their use today should be considered complementary to the experience of doctors and healthcare professionals involved in care. Given the identified limitations, it is evident that both platforms require improvements. The potential for access to more specialized medical databases and continuous algorithmic training promises even more incredible future utility. Ethical considerations cannot be overlooked either. There must be extensive discussion about the quality of information these platforms provide and how user privacy is ensured, as sensitive data may be involved. As these platforms evolve, their utility as diagnostic tools may become more robust, promoting more innovative and secure healthcare applications.

This research paves the way for future investigations into how AI-based tools, like ChatGPT, could be more effectively integrated into clinical practice, offering more reliable and swift diagnoses and potentially enhancing patient outcomes, complementing the human experience.

Our study contributes to a broader conversation about the evolving role of technology in healthcare, specifically in the field of urology, where accurate and timely diagnosis is often critical to treatment success and patient well-being.

## CONCLUSION

We can demonstrate that ChatGPT 3.5 exhibited superior diagnostic accuracy compared to Google in both common and rare urological scenarios. ChatGPT 3.5 displayed acceptable accuracy in habitual conditions cases but was still relatively limited in rare cases. Such findings allow us to glimpse some use of these tools in educational and training processes. Access to medical databases and ongoing development can bring considerable advances, enabling even more robust, innovative, secure, and possibly assisting us in caring for people.

## Data Availability

All data produced in the present work are contained in the manuscript

## Supplementary file 1: Clinical Cases

**Table 1:**
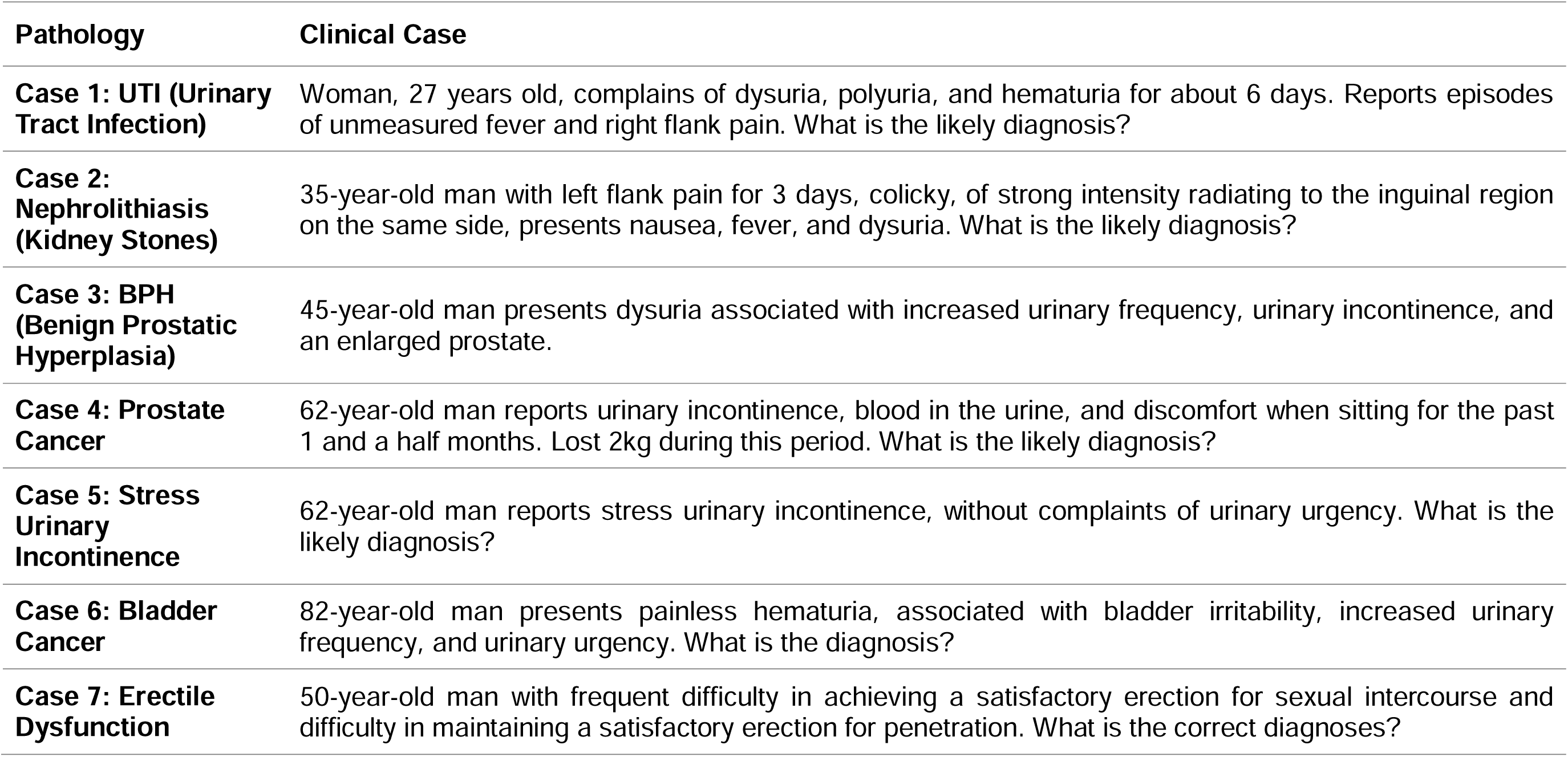
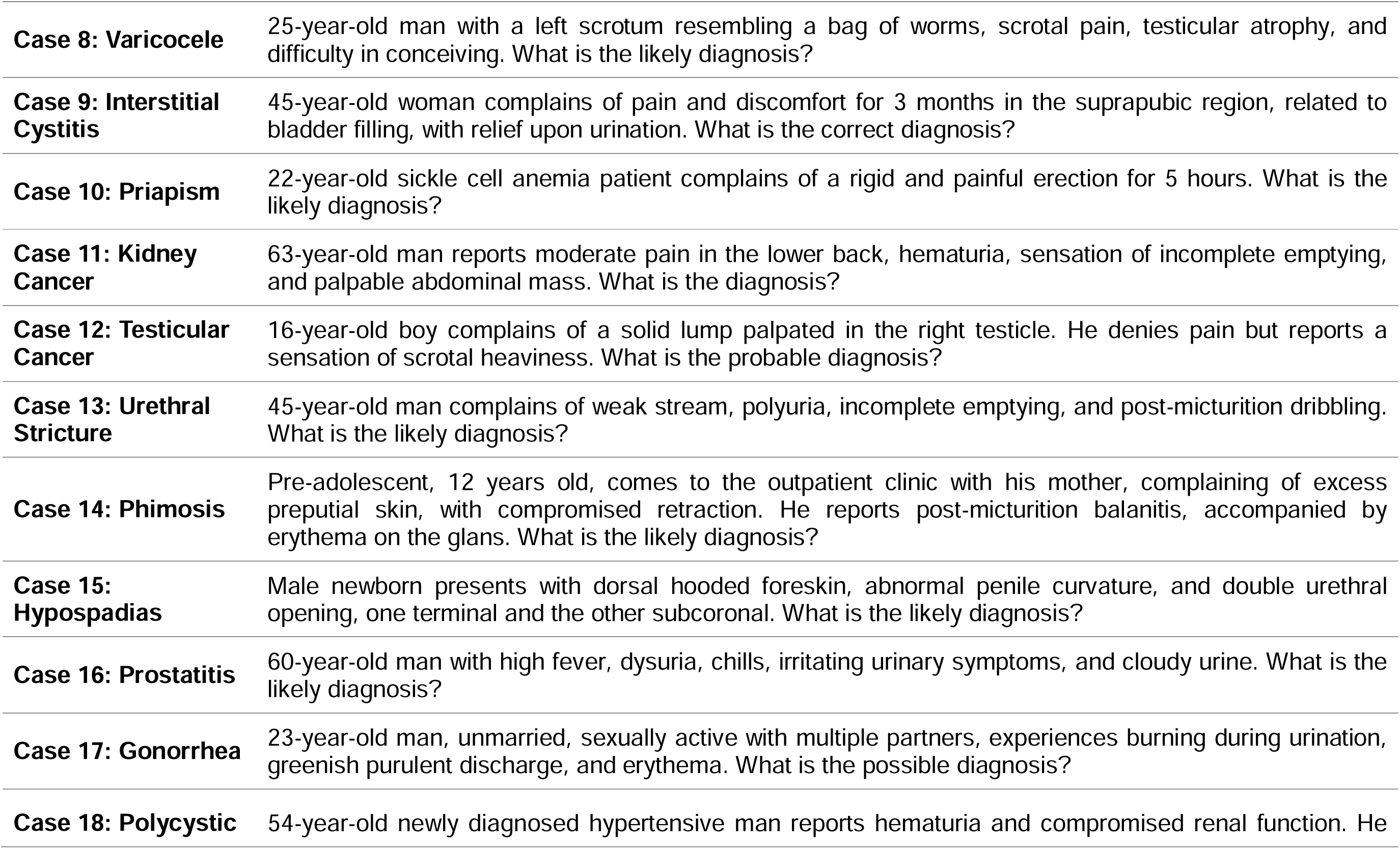
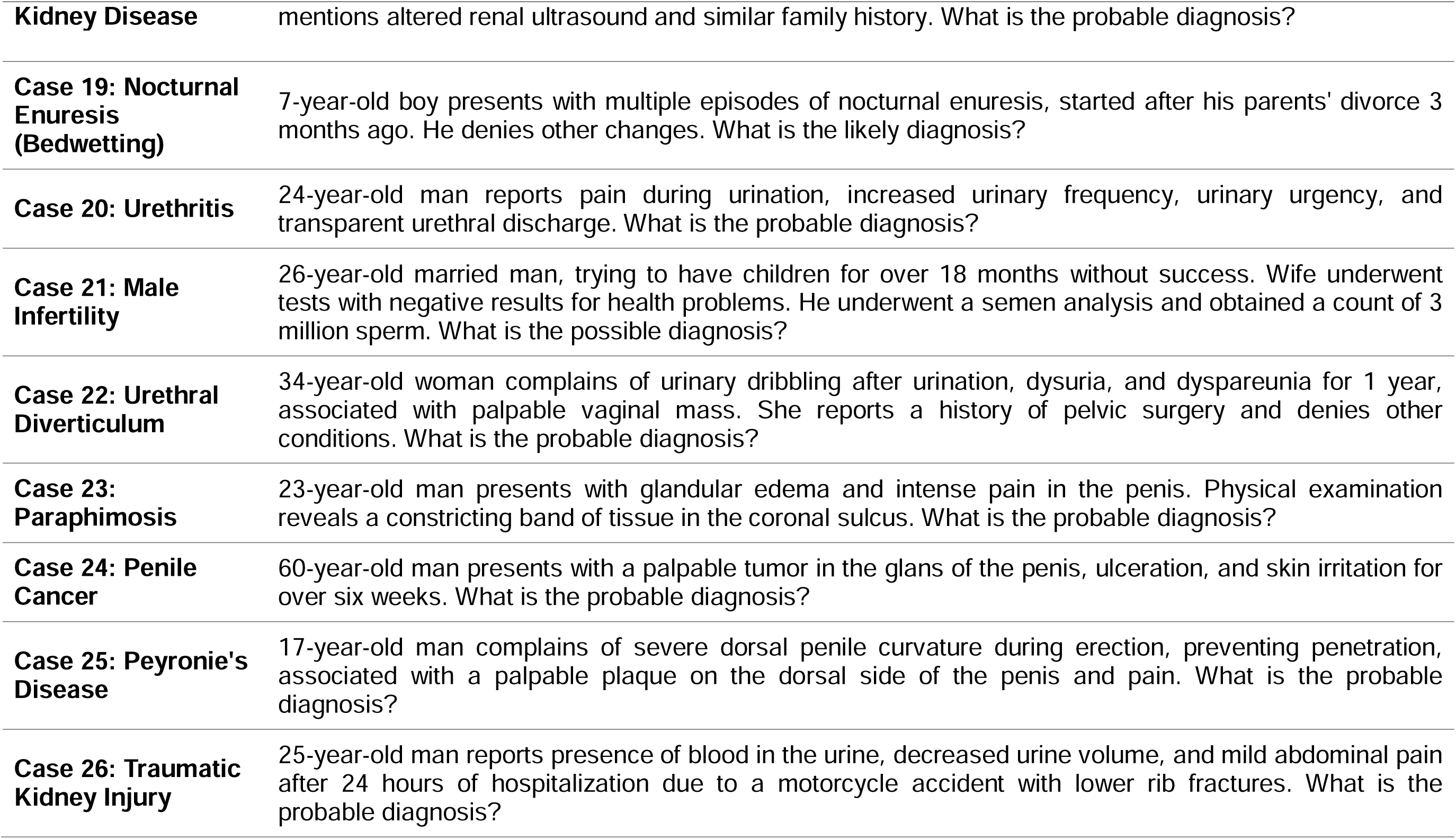
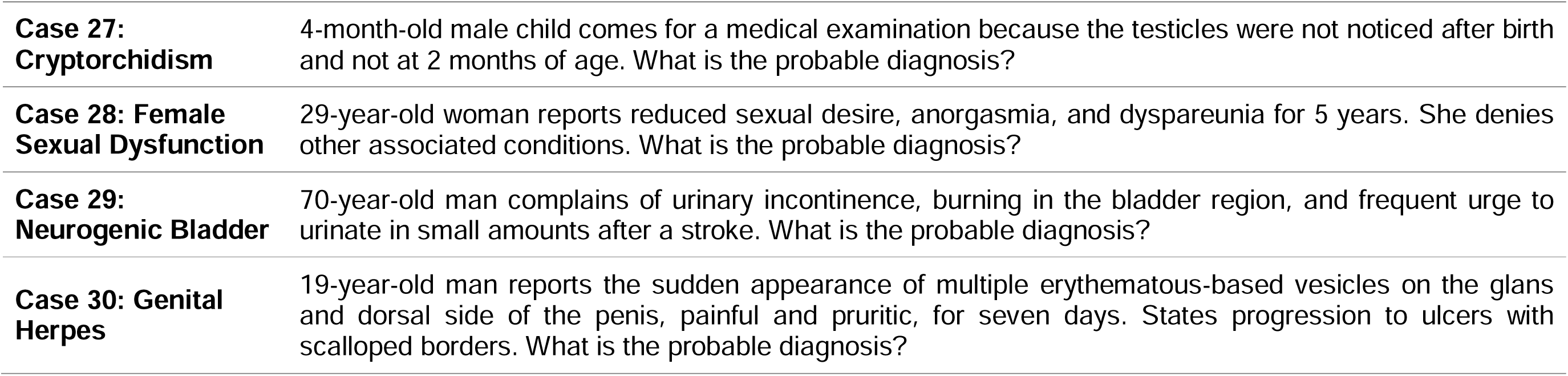
Common Urological Conditions.

**Table 2:**
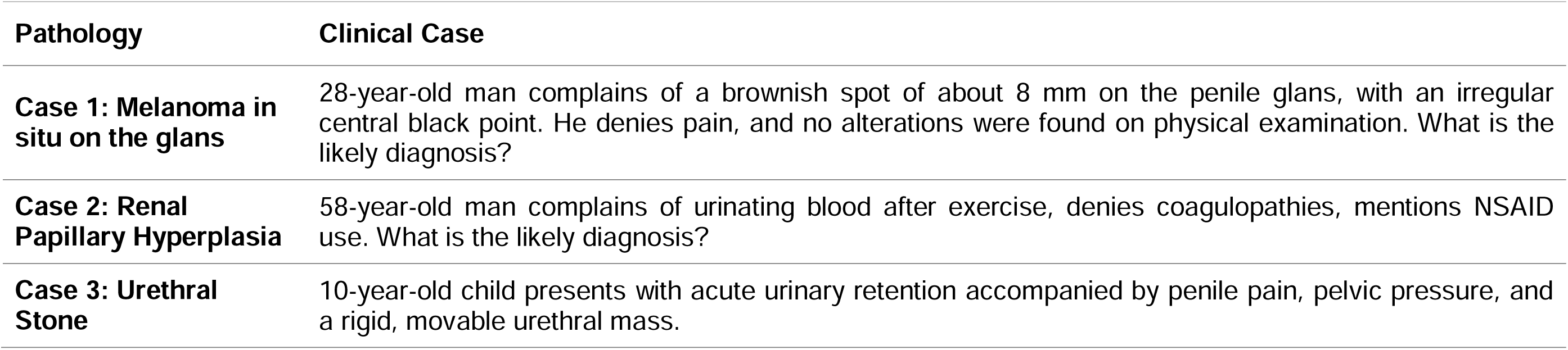
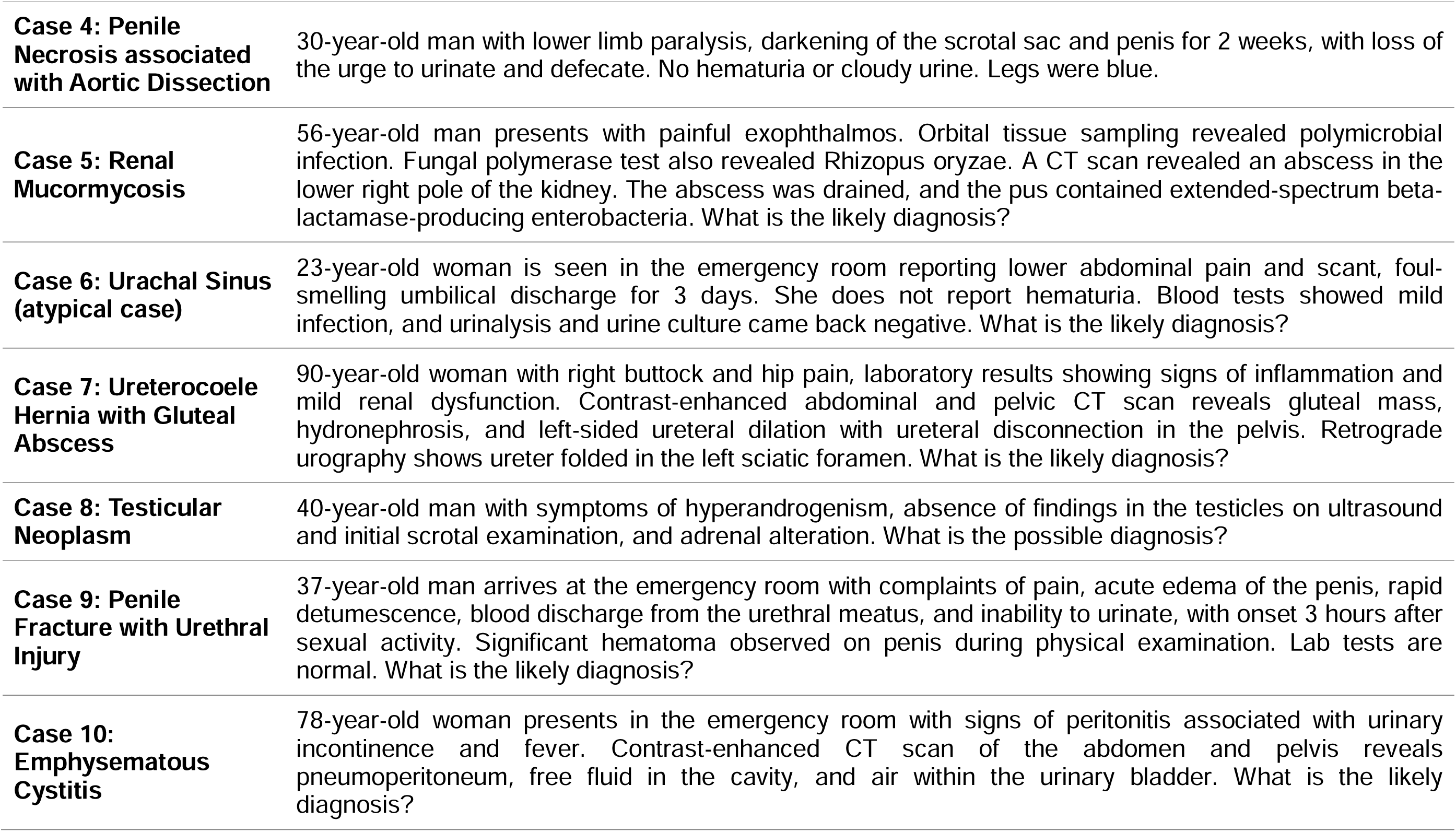
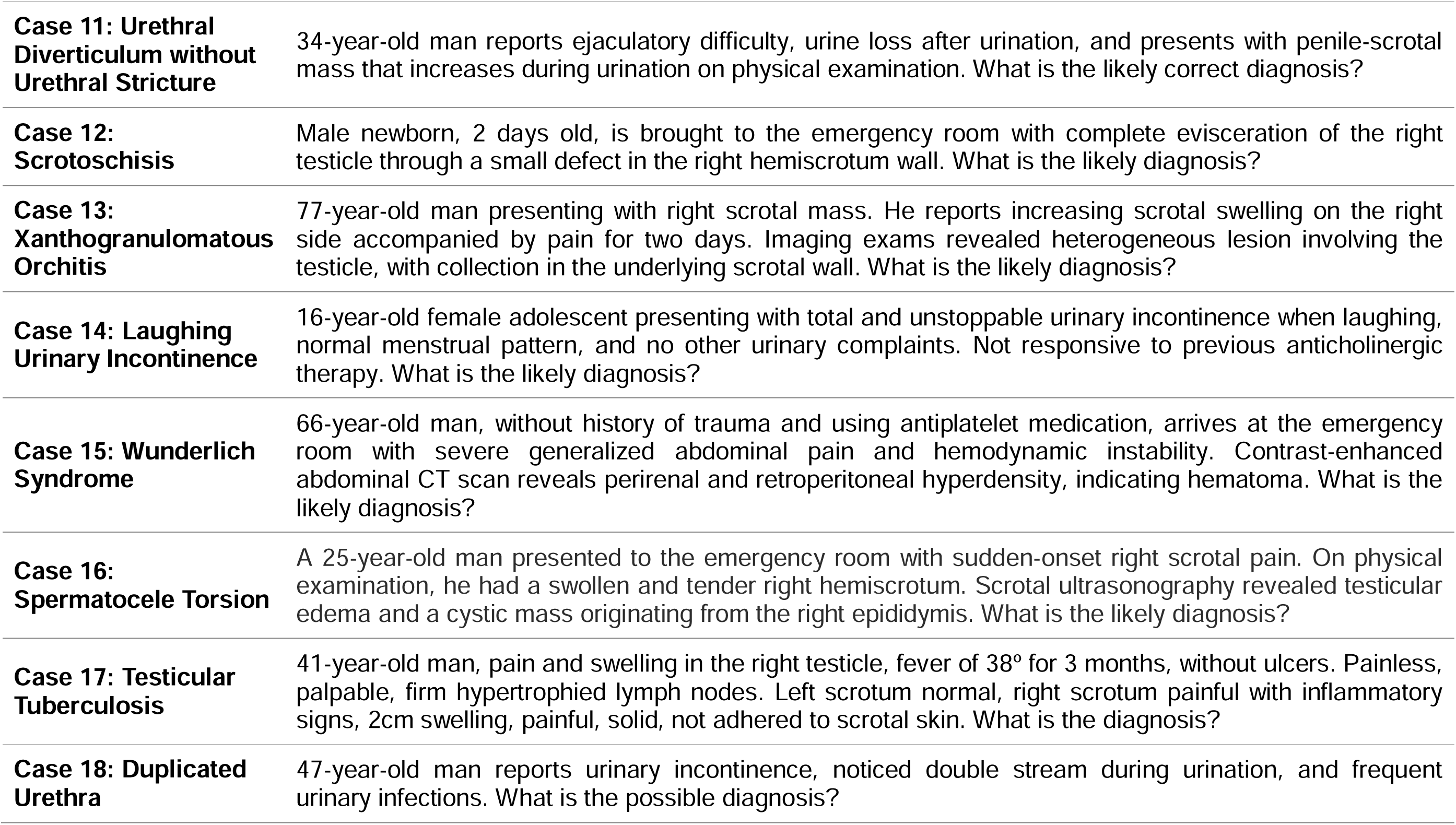
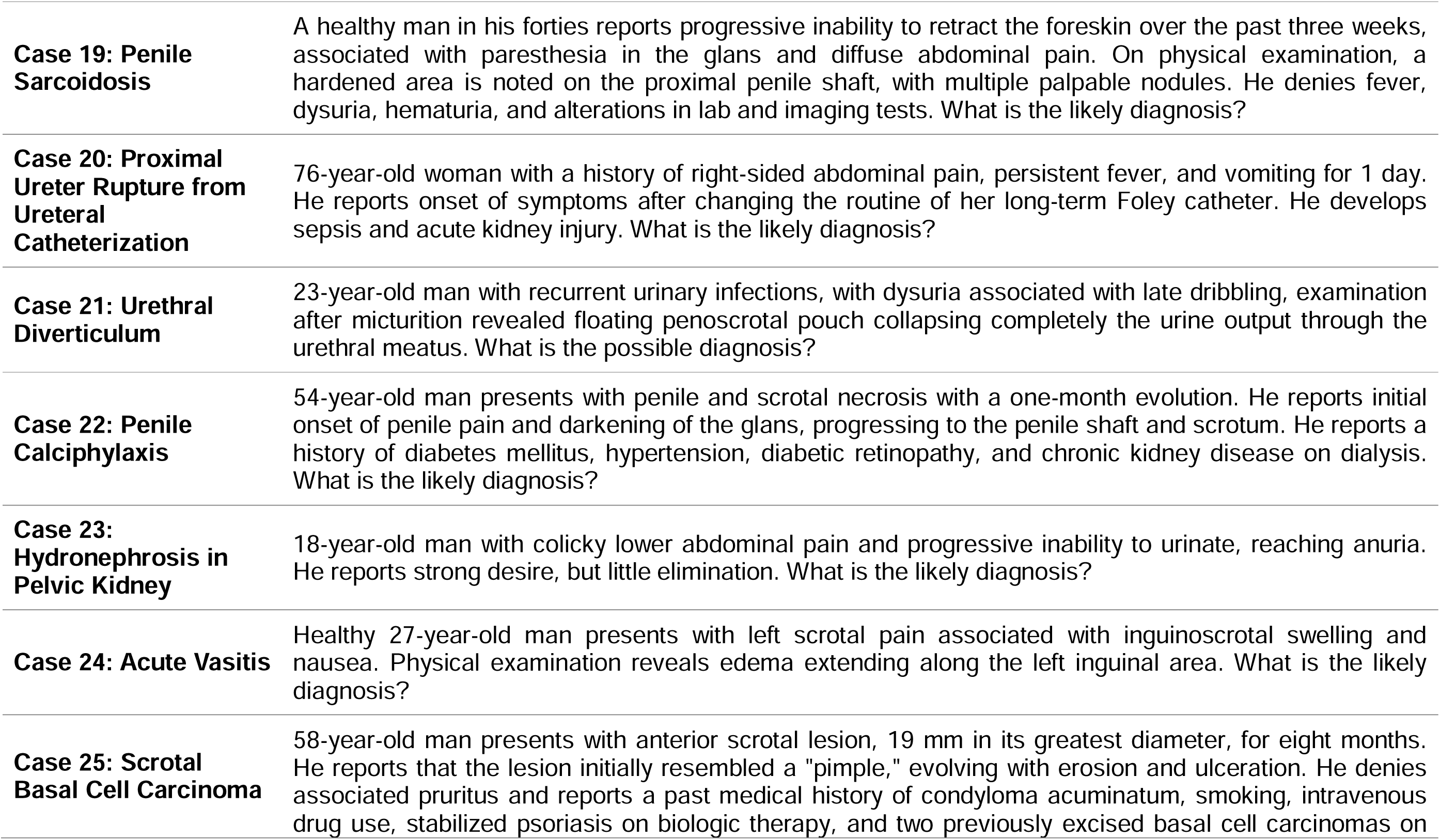
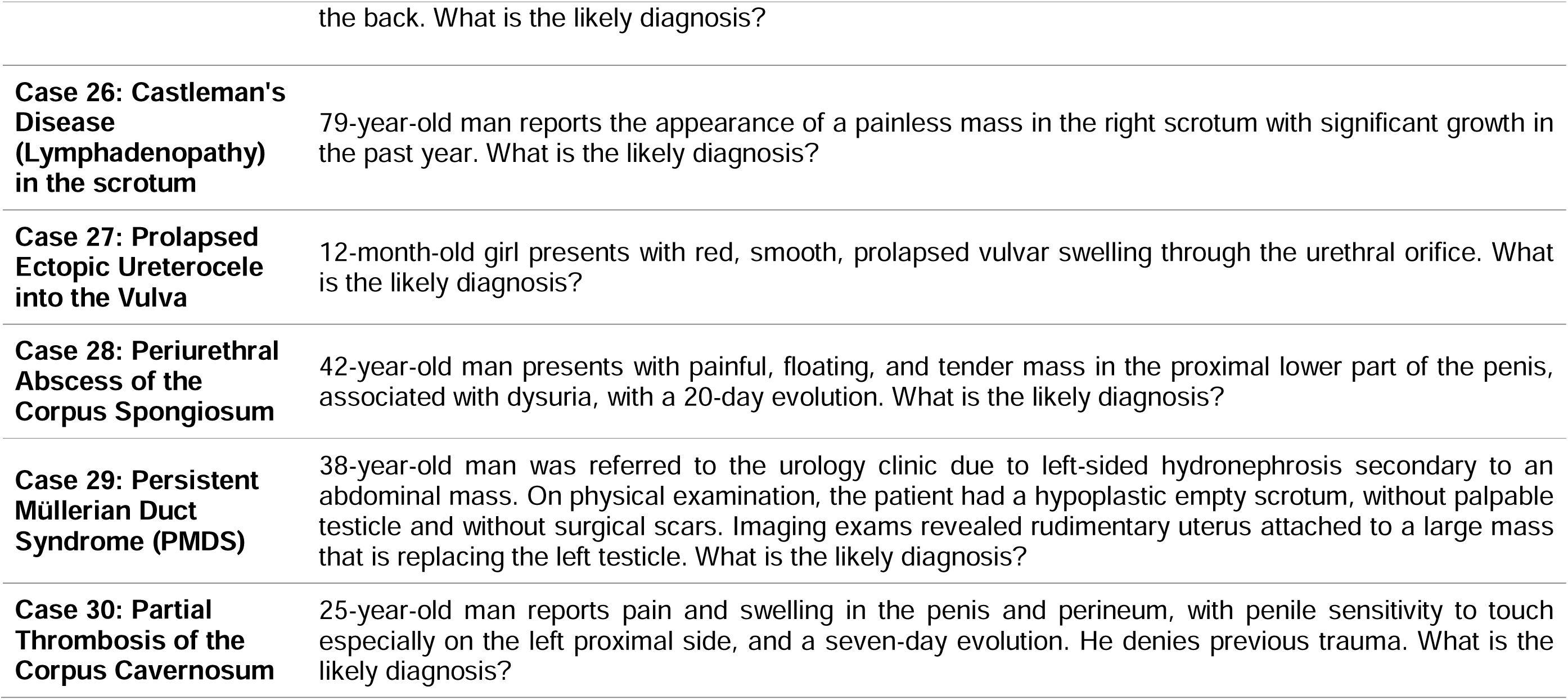
Rare Urological Conditions.

