## Supplementary material for "Diagnosis in Bytes: Comparing the Diagnostic Accuracy of Google and ChatGPT 3.5 as Diagnostic Support Tools": Suplemmentary File 1

**Supplementary file 1: Clinical Cases**

| Table 1: Common Urological Conditions | |
| --- | --- |
| Pathology | **Clinical Case** |
| Case 1: UTI (Urinary Tract Infection) | Woman, 27 years old, complains of dysuria, polyuria, and hematuria for about 6 days. Reports episodes of unmeasured fever and right flank pain. What is the likely diagnosis? |
| Case 2: Nephrolithiasis (Kidney Stones) | 35-year-old man with left flank pain for 3 days, colicky, of strong intensity radiating to the inguinal region on the same side, presents nausea, fever, and dysuria. What is the likely diagnosis? |
| Case 3: BPH (Benign Prostatic Hyperplasia) | 45-year-old man presents dysuria associated with increased urinary frequency, urinary incontinence, and an enlarged prostate. |
| Case 4: Prostate Cancer | 62-year-old man reports urinary incontinence, blood in the urine, and discomfort when sitting for the past 1 and a half months. Lost 2kg during this period. What is the likely diagnosis? |
| Case 5: Stress Urinary Incontinence | 62-year-old man reports stress urinary incontinence, without complaints of urinary urgency. What is the likely diagnosis? |
| Case 6: Bladder Cancer | 82-year-old man presents painless hematuria, associated with bladder irritability, increased urinary frequency, and urinary urgency. What is the diagnosis? |
| Case 7: Erectile Dysfunction | 50-year-old man with frequent difficulty in achieving a satisfactory erection for sexual intercourse and difficulty in maintaining a satisfactory erection for penetration. What is the likely diagnosis? |
| Case 8: Varicocele | 25-year-old man with a left scrotum resembling a bag of worms, scrotal pain, testicular atrophy, and difficulty in conceiving. What is the likely diagnosis? |
| Case 9: Interstitial Cystitis | 45-year-old woman complains of pain and discomfort for 3 months in the suprapubic region, related to bladder filling, with relief upon urination. What is the correct diagnosis? |
| Case 10: Priapism | 22-year-old sickle cell anemia patient complains of a rigid and painful erection for 5 hours. What is the likely diagnosis? |
| Case 11: Kidney Cancer | 63-year-old man reports moderate pain in the lower back, hematuria, sensation of incomplete emptying, and palpable abdominal mass. What is the diagnosis? |
| Case 12: Testicular Cancer | 16-year-old boy complains of a solid lump palpated in the right testicle. He denies pain but reports a sensation of scrotal heaviness. What is the probable diagnosis? |
| Case 13: Urethral Stricture | 45-year-old man complains of weak stream, polyuria, incomplete emptying, and post-micturition dribbling. What is the likely diagnosis? |
| Case 14: Phimosis | Pre-adolescent, 12 years old, comes to the outpatient clinic with his mother, complaining of excess preputial skin, with compromised retraction. He reports post-micturition balanitis, accompanied by erythema on the glans. What is the likely diagnosis? |
| Case 15: Hypospadias | Male newborn presents with dorsal hooded foreskin, abnormal penile curvature, and double urethral opening, one terminal and the other subcoronal. What is the likely diagnosis? |
| Case 16: Prostatitis | 60-year-old man with high fever, dysuria, chills, irritating urinary symptoms, and cloudy urine. What is the likely diagnosis? |
| Case 17: Gonorrhea | 23-year-old man, unmarried, sexually active with multiple partners, experiences burning during urination, greenish purulent discharge, and erythema. What is the possible diagnosis? |
| Case 18: Polycystic Kidney Disease | 54-year-old newly diagnosed hypertensive man reports hematuria and compromised renal function. He mentions altered renal ultrasound and similar family history. What is the probable diagnosis? |
| Case 19: Nocturnal Enuresis (Bedwetting) | 7-year-old boy presents with multiple episodes of nocturnal enuresis, started after his parents' divorce 3 months ago. He denies other changes. What is the likely diagnosis? |
| Case 20: Urethritis | 24-year-old man reports pain during urination, increased urinary frequency, urinary urgency, and transparent urethral discharge. What is the probable diagnosis? |
| Case 21: Male Infertility | 26-year-old married man, trying to have children for over 18 months without success. Wife underwent tests with negative results for health problems. He underwent a semen analysis and obtained a count of 3 million sperm. What is the possible diagnosis? |
| Case 22: Urethral Diverticulum | 34-year-old woman complains of urinary dribbling after urination, dysuria, and dyspareunia for 1 year, associated with palpable vaginal mass. She reports a history of pelvic surgery and denies other conditions. What is the probable diagnosis? |
| Case 23: Paraphimosis | 23-year-old man presents with glandular edema and intense pain in the penis. Physical examination reveals a constricting band of tissue in the coronal sulcus. What is the probable diagnosis? |
| Case 24: Penile Cancer | 60-year-old man presents with a palpable tumor in the glans of the penis, ulceration, and skin irritation for over six weeks. What is the probable diagnosis? |
| Case 25: Peyronie's Disease | 17-year-old man complains of severe dorsal penile curvature during erection, preventing penetration, associated with a palpable plaque on the dorsal side of the penis and pain. What is the probable diagnosis? |
| Case 26: Traumatic Kidney Injury | 25-year-old man reports presence of blood in the urine, decreased urine volume, and mild abdominal pain after 24 hours of hospitalization due to a motorcycle accident with lower rib fractures. What is the probable diagnosis? |
| Case 27: Cryptorchidism | 4-month-old male child comes for a medical examination because the testicles were not noticed after birth and not at 2 months of age. What is the probable diagnosis? |
| Case 28: Female Sexual Dysfunction | 29-year-old woman reports reduced sexual desire, anorgasmia, and dyspareunia for 5 years. She denies other associated conditions. What is the probable diagnosis? |
| Case 29: Neurogenic Bladder | 70-year-old man complains of urinary incontinence, burning in the bladder region, and frequent urge to urinate in small amounts after a stroke. What is the probable diagnosis? |
| Case 30: Genital Herpes | 19-year-old man reports the sudden appearance of multiple erythematous-based vesicles on the glans and dorsal side of the penis, painful and pruritic, for seven days. States progression to ulcers with scalloped borders. What is the probable diagnosis? |

| Table 2: Rare Urological Conditions | |
| --- | --- |
| Pathology | **Clinical Case** |
| Case 1: Melanoma in situ on the glans | 28-year-old man complains of a brownish spot of about 8 mm on the penile glans, with an irregular central black point. He denies pain, and no alterations were found on physical examination. What is the likely diagnosis? |
| Case 2: Renal Papillary Hyperplasia | 58-year-old man complains of urinating blood after exercise, denies coagulopathies, mentions NSAID use. What is the likely diagnosis? |
| Case 3: Urethral Stone | 10-year-old child presents with acute urinary retention accompanied by penile pain, pelvic pressure, and a rigid, movable urethral mass. |
| Case 4: Penile Necrosis associated with Aortic Dissection | 30-year-old man with lower limb paralysis, darkening of the scrotal sac and penis for 2 weeks, with loss of the urge to urinate and defecate. No hematuria or cloudy urine. Legs were blue. |
| Case 5: Renal Mucormycosis | 56-year-old man presents with painful exophthalmos. Orbital tissue sampling revealed polymicrobial infection. Fungal polymerase test also revealed Rhizopus oryzae. A CT scan revealed an abscess in the lower right pole of the kidney. The abscess was drained, and the pus contained extended-spectrum beta-lactamase-producing enterobacteria. What is the likely diagnosis? |
| Case 6: Urachal Sinus (atypical case) | 23-year-old woman is seen in the emergency room reporting lower abdominal pain and scant, foul-smelling umbilical discharge for 3 days. She does not report hematuria. Blood tests showed mild infection, and urinalysis and urine culture came back negative. What is the likely diagnosis? |
| Case 7: Ureterocoele Hernia with Gluteal Abscess | 90-year-old woman with right buttock and hip pain, laboratory results showing signs of inflammation and mild renal dysfunction. Contrast-enhanced abdominal and pelvic CT scan reveals gluteal mass, hydronephrosis, and left-sided ureteral dilation with ureteral disconnection in the pelvis. Retrograde urography shows ureter folded in the left sciatic foramen. What is the likely diagnosis? |
| Case 8: Testicular Neoplasm | 40-year-old man with symptoms of hyperandrogenism, absence of findings in the testicles on ultrasound and initial scrotal examination, and adrenal alteration. What is the possible diagnosis? |
| Case 9: Penile Fracture with Urethral Injury | 37-year-old man arrives at the emergency room with complaints of pain, acute edema of the penis, rapid detumescence, blood discharge from the urethral meatus, and inability to urinate, with onset 3 hours after sexual activity. Significant hematoma observed on penis during physical examination. Lab tests are normal. What is the likely diagnosis? |
| Case 10: Emphysematous Cystitis | 78-year-old woman presents in the emergency room with signs of peritonitis associated with urinary incontinence and fever. Contrast-enhanced CT scan of the abdomen and pelvis reveals pneumoperitoneum, free fluid in the cavity, and air within the urinary bladder. What is the likely diagnosis? |
| Case 11: Urethral Diverticulum without Urethral Stricture | 34-year-old man reports ejaculatory difficulty, urine loss after urination, and presents with penile-scrotal mass that increases during urination on physical examination. What is the likely correct diagnosis? |
| Case 12: Scrotoschisis | Male newborn, 2 days old, is brought to the emergency room with complete evisceration of the right testicle through a small defect in the right hemiscrotum wall. What is the likely diagnosis? |
| Case 13: Xanthogranulomatous Orchitis | 77-year-old man presenting with right scrotal mass. He reports increasing scrotal swelling on the right side accompanied by pain for two days. Imaging exams revealed heterogeneous lesion involving the testicle, with collection in the underlying scrotal wall. What is the likely diagnosis? |
| Case 14: Laughing Urinary Incontinence | 16-year-old female adolescent presenting with total and unstoppable urinary incontinence when laughing, normal menstrual pattern, and no other urinary complaints. Not responsive to previous anticholinergic therapy. What is the likely diagnosis? |
| Case 15: Wunderlich Syndrome | 66-year-old man, without history of trauma and using antiplatelet medication, arrives at the emergency room with severe generalized abdominal pain and hemodynamic instability. Contrast-enhanced abdominal CT scan reveals perirenal and retroperitoneal hyperdensity, indicating hematoma. What is the likely diagnosis? |
| Case 16: Spermatocele Torsion | A 25-year-old man presented to the emergency room with sudden-onset right scrotal pain. On physical examination, he had a swollen and tender right hemiscrotum. Scrotal ultrasonography revealed testicular edema and a cystic mass originating from the right epididymis. What is the likely diagnosis? |
| Case 17: Testicular Tuberculosis | 41-year-old man, pain and swelling in the right testicle, fever of 38º for 3 months, without ulcers. Painless, palpable, firm hypertrophied lymph nodes. Left scrotum normal, right scrotum painful with inflammatory signs, 2cm swelling, painful, solid, not adhered to scrotal skin. What is the diagnosis? |
| Case 18: Duplicated Urethra | 47-year-old man reports urinary incontinence, noticed double stream during urination, and frequent urinary infections. What is the possible diagnosis? |
| Case 19: Penile Sarcoidosis | A healthy man in his forties reports progressive inability to retract the foreskin over the past three weeks, associated with paresthesia in the glans and diffuse abdominal pain. On physical examination, a hardened area is noted on the proximal penile shaft, with multiple palpable nodules. He denies fever, dysuria, hematuria, and alterations in lab and imaging tests. What is the likely diagnosis? |
| Case 20: Proximal Ureter Rupture from Ureteral Catheterization | 76-year-old woman with a history of right-sided abdominal pain, persistent fever, and vomiting for 1 day. He reports onset of symptoms after changing the routine of her long-term Foley catheter. He develops sepsis and acute kidney injury. What is the likely diagnosis? |
| Case 21: Urethral Diverticulum | 23-year-old man with recurrent urinary infections, with dysuria associated with late dribbling, examination after micturition revealed floating penoscrotal pouch collapsing completely the urine output through the urethral meatus. What is the possible diagnosis? |
| Case 22: Penile Calciphylaxis | 54-year-old man presents with penile and scrotal necrosis with a one-month evolution. He reports initial onset of penile pain and darkening of the glans, progressing to the penile shaft and scrotum. He reports a history of diabetes mellitus, hypertension, diabetic retinopathy, and chronic kidney disease on dialysis. What is the likely diagnosis? |
| Case 23: Hydronephrosis in Pelvic Kidney | 18-year-old man with colicky lower abdominal pain and progressive inability to urinate, reaching anuria. He reports strong desire, but little elimination. What is the likely diagnosis? |
| Case 24: Acute Vasitis | Healthy 27-year-old man presents with left scrotal pain associated with inguinoscrotal swelling and nausea. Physical examination reveals edema extending along the left inguinal area. What is the likely diagnosis? |
| Case 25: Scrotal Basal Cell Carcinoma | 58-year-old man presents with anterior scrotal lesion, 19 mm in its greatest diameter, for eight months. He reports that the lesion initially resembled a "pimple," evolving with erosion and ulceration. He denies associated pruritus and reports a past medical history of condyloma acuminatum, smoking, intravenous drug use, stabilized psoriasis on biologic therapy, and two previously excised basal cell carcinomas on the back. What is the likely diagnosis? |
| Case 26: Castleman's Disease (Lymphadenopathy) in the scrotum | 79-year-old man reports the appearance of a painless mass in the right scrotum with significant growth in the past year. What is the likely diagnosis? |
| Case 27: Prolapsed Ectopic Ureterocele into the Vulva | 12-month-old girl presents with red, smooth, prolapsed vulvar swelling through the urethral orifice. What is the likely diagnosis? |
| Case 28: Periurethral Abscess of the Corpus Spongiosum | 42-year-old man presents with painful, floating, and tender mass in the proximal lower part of the penis, associated with dysuria, with a 20-day evolution. What is the likely diagnosis? |
| Case 29: Persistent Müllerian Duct Syndrome (PMDS) | 38-year-old man was referred to the urology clinic due to left-sided hydronephrosis secondary to an abdominal mass. On physical examination, the patient had a hypoplastic empty scrotum, without palpable testicle and without surgical scars. Imaging exams revealed rudimentary uterus attached to a large mass that is replacing the left testicle. What is the likely diagnosis? |
| Case 30: Partial Thrombosis of the Corpus Cavernosum | 25-year-old man reports pain and swelling in the penis and perineum, with penile sensitivity to touch especially on the left proximal side, and a seven-day evolution. He denies previous trauma. What is the likely diagnosis? |
